# Good work in the COVID-19 recovery: priorities and changes for the future

**DOI:** 10.1101/2022.01.31.22270163

**Authors:** Melda Lois Griffiths, Benjamin J Gray, Richard G Kyle, Alisha R Davies

## Abstract

Employment is a wider determinant of health, and the COVID-19 pandemic has disrupted working lives, with individuals having to adapt to new ways of working. These new experiences may shape what kind of work people want in future. This research used a sample of working adults in Wales to identify the workforce’s priorities for future work, and the employment changes that they have considered making since the start of the COVID-19 pandemic. Data was collected at two time-points (May-June 2020; December 2020-January 2021) in a nationally-representative longitudinal household survey across Wales.

Work priorities remained largely stable throughout the pandemic, however the desire to work close to home increased as the pandemic progressed. Those in poorer health prioritised flexibility, and were more likely to consider retiring than their healthier counterparts. Becoming self-employed was more likely to be considered by those with limiting pre-existing conditions or low mental well-being. Over 20% of the total sample had considered retraining, with those with low mental well-being, younger individuals and those experiencing financial insecurity being more likely to consider doing so. Furloughed individuals were more likely to consider retraining, becoming self-employed, securing permanent employment and compressing their working hours.

Those prone to facing insecurity within their working lives (those that were furloughed, those experiencing financial insecurity, and those in ill-health) were all more likely to consider changing their employment conditions – these groups may require additional support in accessing secure and flexible work. Action is needed to ensure that good work, that is good for health, is equally accessible for all.

## Introduction

As a wider determinant of health, employment can both positively and negatively affect our health and quality of life (Burgard and Lin 2013; van Aerden et al. 2016; Julià et al. 2017). Good work, that is stable, meaningful and fairly compensated, is known to be good for health (Waddell and Burton 2006). Enabling individuals to have access to what constitutes good work for them and their circumstances is vital to ensure equitable access to healthy working lives for all.

Good, fair work has seen policy-level support both internationally and within the UK (e.g. European Parliament’s employment package (European Parliament 2012), UK Government’s Good Work Plan (UK Government 2018), and Welsh Government’s Employability Plan and Fair Work Wales report (Welsh Government 2018, 2019)). These strategies included elements such as ensuring good quality, fairly-rewarded, flexible and secure work, and supporting lifelong learning and skill development. With the policy-landscape acknowledging the importance of job quality, and prioritising various aspects of work (such as pay, security and flexibility), the development of insights that shed light on the priorities and intentions of the workforce itself will help ensure alignment between policy and workforce-needs. Specific groups of the population may face different barriers to accessing employment (Welsh Government 2018) or have different priorities for work. Capturing how these translate to priorities for future work, or intentions for future employment is necessary.

This is more so true within the context of the COVID-19 pandemic recovery. The pandemic has had a disruptive influence on the world of work, and required individuals to rapidly adapt to new ways of working e.g. working from home, in-work changes, furlough (Daniel 2020; Kramer and Kramer 2020; Leone et al. 2020; Galanti et al. 2021; Szulc and Smith 2021). Some elements within the policy-level intentions outlined above were resultantly forced into fruition e.g. the need for more flexible working arrangements and working from home. On the other hand, others became more difficult to achieve e.g. those that were furloughed or became unemployed experienced more insecurity.

While these dramatic changes to the population’s employment related experiences were welcomed by some (e.g. those enjoying greater flexibility through home-working), they led to increased isolation or financial strain for others (Dyakova et al. 2021; Green et al. 2021). Furthermore, evidence has shown that population groups that already face health inequities were disproportionately affected by the pandemic’s negative impacts, exacerbating pre-existing societal inequalities (Gray et al. 2021). For example, the youngest and eldest in society, along with those with less financial security were more likely to be furloughed, and those with non-permanent employment contracts, low mental well-being or household financial difficulties were more likely to become unemployed (Gardiner and Slaughter 2020; Mayhew and Anand 2020; Crossley et al. 2021; Gray et al. 2021). Resulting uncertainty and increased financial insecurity may have spurred individuals to reconsider their current employment conditions and explore alternative options for the future. While this dichotomy of work-related experiences arose in response to the pandemic, they could have produced shifts in the public’s priorities and intentions for future work which might have longstanding societal and policy-level implications beyond the pandemic itself (Kramer and Kramer 2020). Shedding light on these priorities and intentions, and how they may have changed during the pandemic, will help inform the direction of future policies that support good, fair work.

This study therefore firstly aimed to establish the employment priorities of employed Welsh working age adults at two time-points within the COVID-19 pandemic. Secondly, the study aimed to capture the employment changes that these individuals had considered making since the pandemic began. For both, comparisons were made across socio-demographic groups, employment and income, and health status.

## Methods

### Study design

A nationally-representative longitudinal household survey was undertaken across Wales (*COVID-19, Employment and Health in Wales* study) with a paper-to-web push approach. The Health Research Authority approved the study (IRAS: 282223). Data was collected at two time-points - T1 data collection occurred between May-June 2020, and the follow-up at T2 between November 2020 and January 2021.

### Study population and recruitment

All working age adults aged between 16-64 years resident in Wales, in current employment as of February 2020, were eligible, with those in full-time education or unemployed being excluded. To obtain a sample that was representative of the Welsh population, a stratified random probability sampling framework by age, gender and deprivation quintile was used. Respondents were informed that their participation was voluntary and that their responses would be confidential. Reminder letters were sent 10 days following original invitation. For each household, the eligible adult with the next birthday was asked to participate. A total of 1,382 adults responded at T1 (7.0% response rate), with 1,019 being from within the main sample (7.0% response rate), and 273 from the booster sample (5.5% response rate). Full details of the recruitment and sampling strategy are discussed elsewhere (Gray et al, 2021). Of the 1,382 adults that responded to the initial survey at T1, 1,084 individuals gave permission to be contacted for a follow up study. For these individuals, the follow-up data collection phase was from November 2020 to January 2021. If a valid email address was provided (N=925), individuals were emailed an invitation to take part a second time with two further email remainders to encourage participation. If a valid email address was not provided (N=159), individuals were sent a postal invitation and one reminder invitation. In total, 626 individuals completed the follow-up online questionnaire at T2 (58% response rate). Nine responses were excluded as identification codes were inputted incorrectly, leaving a sample of 615 (98.2%). To allow for longitudinal comparisons, this study uses the responses of this sample of 615 individuals who provided observations at both T1 and T2.

### Questionnaire measures

Questionnaire measures for the two dependent variables (employment priorities and considered changes) can be seen in Supplementary Material 1. At T1 and T2, respondents were asked to indicate their five greatest priorities for any new or future work from the following options: having a workplace close to home; flexible working conditions; opportunities for personal/professional development; availability of childcare; reliable local transport services; pay package (including salary, pension and benefits); hours of work; how interesting, enjoyable or rewarding the work is; how well the job matches qualifications, skills and experiences; and job security. At T2, respondents were asked an additional question - which employment changes had they considered making since the start of the COVID-19 pandemic (February 2020)? Options were as follows: retraining to do a different job; upskilling for a promotion; securing a permanent contract; compressing working hours; going part-time; becoming self-employed/freelance; retiring; or none of the above.

To explore the extent to which work priorities and considered changes differed across population groups, measurements from questions relating to socio-economic status, health and employment/income were also used to build a logistic regression model. Explanatory variables included age group, gender, deprivation quintile (assigned using the Welsh Index of Multiple Deprivation (Welsh Government 2021a) from residential postcode), individual self-reported general health and presence of limiting pre-existing conditions (using validated questions from the National Survey for Wales (Welsh Government 2021b)), and mental well-being (using the shortened Warwick Edinburgh Mental Well-being Scale (Stewart-Brown et al. 2011) and using 1 SD below the mean as our cut-off score for low mental well-being). Explanatory variables relating to employment and income were also adopted, including employment contract type (permanent, fixed term, atypical, self-employed/freelance), furlough status, wage precariousness to explore financial insecurity (computed across three variables (see Supplementary Material 1) and based on the Employment Precariousness Scale (Vives et al. 2015)) and job skill level (calculated using the Standard Occupational Classification for the UK (Office for National Statistics 2020a)).

### Statistical approach

Statistical analysis was undertaken in IBM SPSS Statistics (Version 24). Chi-square (χ^2^ and Fisher’s Exact tests were used to explore associations across socio-demographic groups, employment and income, and health status to provide insights into which components of work different sub-groups considered as priorities, and the employment changes that different groups had considered making. Multivariate logistic regressions were used to identify independent predictors of employment priorities and considered changes, including socio-economic, employment and income, and health factors as described above. Selected longitudinal comparisons were made using McNemar’s tests.

## Results

### Sample Characteristics

Respondents predominantly identified as women (63.7% compared to 35.4% men). The sample was biased towards those between 40 and 59 years of age (40-49 25%; 50-59 33.3%), however there was representation across all working ages. Respondents were well distributed across deprivation quintiles (see Supplementary Material 2 for full sample characteristics).

### Prioritised components of work (Aim 1)

As shown in Figure 1, at both time-points, the five components of work prioritised by the majority of respondents were pay (T1: 76.4%; T2: 76.4%); how interesting, enjoyable or rewarding the work was (T1: 68.4%; T2: 67.6%); hours of work (T1: 62.8%; T2: 62%); how close the workplace was to where individuals lived (T1: 56%; T2: 62%) and flexible working conditions (T1: 54.7%; T2: 55.2%). Half the sample also prioritised job security at T1, but this decreased below half at T2 (T1: 50.6%; T2: 48.3%). Availability of childcare and reliable local transport were prioritized by less than 10% of the sample at both time-points (see Discussion).

**Figure 1.**
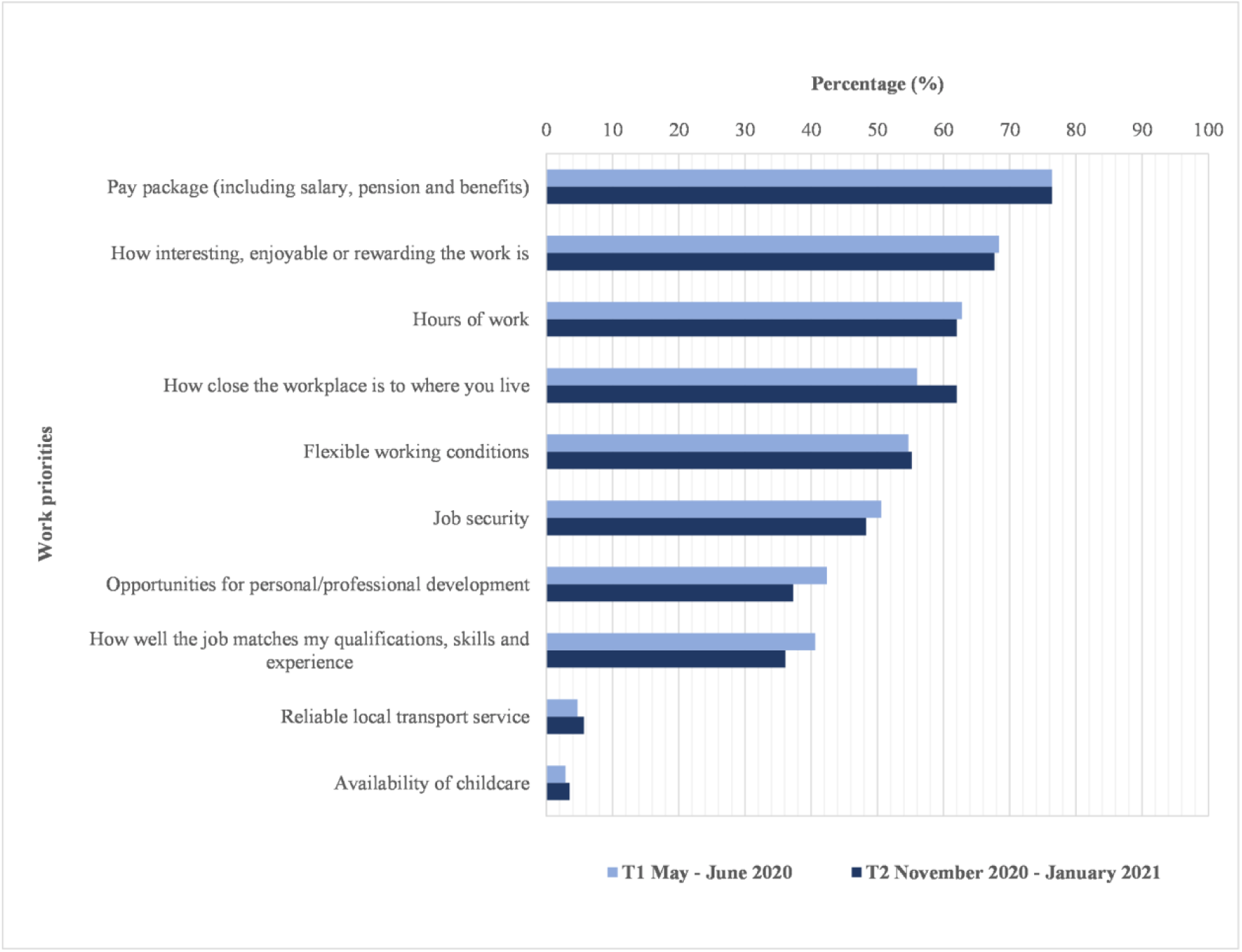
Percentage within sample of Welsh working age adults selecting each priority for future work at two time-points during the COVID-19 pandemic. Respondents were asked to select five from those listed.

The components of work being prioritized remained largely unchanged when comparing T1 and T2 measures, however individuals were more likely to prioritise having a workplace that was close to their home by T2 (+6 percentage points; *p* = .01), and less like to prioritise having work that offered opportunities for development (−4.9 percentage points; *p* = .003)) or work that matched their qualifications, skills or experiences (−4.5 percentage points; *p* = .02). No significant changes were found between T1 and T2 for any other components of work when comparing across the whole sample.

#### Comparison of priorities across characteristics

Comparisons across socio-demographic groups, employment and income, and health status were carried out for the work priorities selected by 50% or more of the sample at any one time-point (leaving six work priorities). For both time-points, the selection of each priority was compared across groups. The percentage selecting a priority, and the associations between factors and the selection of work priorities are documented in full in the supplemental materials (SM1), as are the findings of multivariate logistic regression models that indicated the significant predictors for the selection of each work priority (e.g. gender, age, contract type). Longitudinal effects were explored for factors that were associated with the selection of a work priority at only one of the two time-points.

##### Priorities by socio-economic and employment/income characteristics

*Flexible work* was prioritised by a smaller proportion of younger (under 30) individuals and furloughed individuals at T1, however by T2 they became as likely as their older or non-furloughed counterparts to prioritise flexibility (see SM1). Flexible work was also more likely to be prioritised by individuals with children in their households, with over 60% selecting it as a priority at both time-points (T1: aOR 2.21 [95% CI 1.26-3.89]; T2: aOR 1.76 [95% CI 1.02-3.04]).

*Pay* was less likely to be prioritised by those in atypical or self-employment at both time-points (Atypical T1: aOR 0.28 [95% CI 0.08-0.99]; Atypical T2: aOR 0.16 [95% CI 0.05-0.53]; Self-employed T1: aOR 0.15 [95% CI 0.06-0.33]; Self-employed T2: aOR 0.24 [95% CI 0.11-0.55]). Those with fixed term contracts were also less likely to prioritise pay at T1 (aOR 0.26 [95% CI 0.10-0.71]). Lastly, individuals with high wage precarity were consistently less likely than those with low wage precarity to prioritise their pay (T1: aOR 0.29 [95% CI 0.14-0.59]); T2: aOR 0.35 [95% CI 0.16-0.73]).

*Working hours* were prioritized by a greater proportion of women than men, and a greater proportion of those aged 40 or above than younger respondents at both time-points (see SM1). At T2, those living in the second most deprived areas (WIMD 2) were twice as likely as those living in the least deprived areas to prioritise their working hours (aOR 2.04 [95% CI 1.07-3.87]). At the same time-point, hours were more likely to be prioritized by those with high (aOR 2.45 [95% CI 1.28-4.69]) or moderate wage precarity at T2 (aOR 2.28 [95% CI 1.34-3.86]).

*Working close to home* became increasingly important over time for those on permanent (T1 = 56.5%; T2 = 63.6%, *p*=.02) and fixed term contracts (T1 = 61.5%; T2 = 73.5%, *p*=.002), and for those that were not furloughed (T1 = 53.6%; T2 = 61.9%, *p*=.001). When looking across employment conditions, the self-employed were less likely to prioritise working close to home than their counterparts with permanent contracts (aOR 0.32 [95% CI 0.14-0.71]). Furthermore, at both time-points, those with high as opposed to low wage precariousness were twice as likely to prioritise having a workplace close to home (TI: aOR 2.11 [95% CI 1.14-3.91]; T2: aOR 2.04 [95% CI 1.08-3.87]).

*Job security* was less likely to be prioritised by those that were self-employed with less than 25% placing it as a priority at both time-points (T1: aOR 0.19 [95% CI 0.08-0.43]); T2: aOR 0.22 [95% CI 0.09-0.53]). At T1, those in atypical employment were also less likely to prioritise job security (aOR 0.20 [95% CI 0.05-0.82]). Those with permanent contracts were the most concerned about job security, with over 50% placing it as a priority at both time-points.

*Having enjoyable, interesting or rewarding work* was less likely to be prioritised by those in fixed term (aOR 0.33 [95% CI 0.13-0.88]) or atypical employment (aOR 0.14 [95% CI 0.04-0.53]) at T2. More secure, permanent work was therefore more likely to be associated with prioritising in-work enjoyment. In the same vein, those that experienced less financial insecurity (i.e. low wage precarity) were significantly more likely to prioritise having enjoyable, interesting and rewarding work (over 75% at T1 and T2) than those with high wage precariousness (66.7% at T1 and 56.4% at T2).

##### Priorities by self-reported health characteristics

*Flexible work* was consistently more likely to be prioritised by those in poorer health (T1: aOR 2.06 [95% CI 1.10-3.88]; T2; aOR 1.87 [95% CI 1.05-3.33], with approximately 64% of those that were not in good health placing it as a priority, compared to 51.8% of their healthier counterparts.

*Pay* was more likely to be prioritised by those with low mental well-being at T1 (aOR 4.39 [95% CI 1.62-11.92]). In contrast, those with limiting pre-existing conditions were significantly less likely to prioritise pay (69.5%) when comparing to those without at T2 (78.1%).

*Working close to home* was equally important for those with limiting pre-existing conditions at both time-points (T1 = 64.4%; T2 = 67.2%, *p*=.39), however it became increasingly important to those without limiting pre-existing conditions as time progressed (T1 = 52.6%; T2 = 61.1%; *p*=.02).

*Having enjoyable, interesting or rewarding work* was more likely to be prioritised by those with limiting pre-existing conditions at T1 (aOR 1.97 [95% CI 1.08-3.57]). However, at the same time-point, those with low mental well-being were less likely to prioritise in-work enjoyment at T1 (aOR 0.47 [95% CI 0.24-0.92]).

### Employment changes (Aim 2)

Of the proposed employment changes in the questionnaire, retraining to do a different job, upskilling for a promotion, going part-time and retiring were the changes considered by the highest proportion of respondents (Figure 2). Nearly half of our sample considered not making any of the employment changes listed since the start of the COVID-19 pandemic (275 respondents, 46.7%).

**Figure 2.**
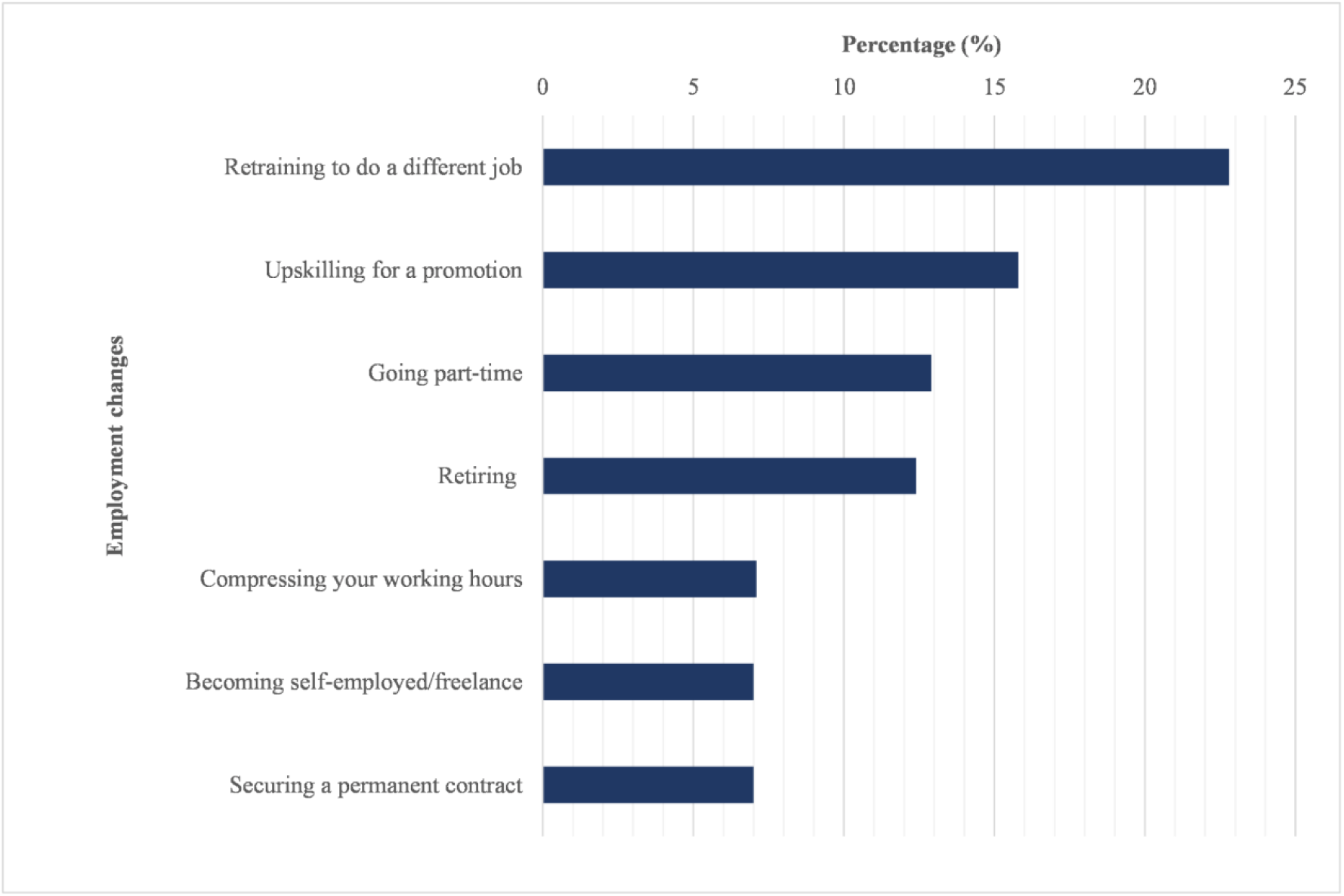
Percentage of Welsh working age adult sample considering each of the employment changes listed at T2. Respondents could select any of the changes listed, note other changes considered, or state that they had not considered making any changes.

#### Consideration of employment changes across groups

The percentage of respondents within various socio-demographic, employment and health groups considering each employment change (or none at all) can be seen in SM2, along with the results of multivariate logistic regression models that identified significant predictors of considering each change.

##### Employment changes by socio-economic and employment/income characteristics

*Retraining* was more likely to be considered by younger age groups (than those aged 50 or above), and less likely to be considered by those in the second and third deprivation quintiles (see SM2). Furloughed individuals were more than twice as likely as non-furloughed individuals to consider retraining (aOR 2.34 [95% CI 1.22-4.49]), as were those indicating high (as opposed to low) wage precarity (aOR 2.25 [95% CI 1.02-4.94]). Half of those with atypical employment contracts had considered retraining, while less than 30% of respondents with all other contract types had done the same.

*Becoming self-employed/freelancing* was more likely to be considered by younger respondents (under 40), with those in their 30s being nearly four times more likely to do so than those in their 40s (aOR 3.79 [95% CI 1.12-12.86]). Furloughed individuals were more than four times more likely to consider becoming self-employed/freelance, compared to their non-furloughed counterparts (aOR 4.64 [95% CI 1.71-12.53]).

*Upskilling for a promotion* was far less likely to be considered by those aged 50 or above when compared with those in their 40s (50-59: aOR 0.20 [95% CI 0.07-0.54]); 60-64: aOR 0.11 [95% CI 0.02-0.55]). In contrast, 38.1% of those under 30 had thought about upskilling, with this age group being three times more likely to do so than those in their 40s (aOR 2.95 [95% CI 1.13-7.71]).

*Securing permanent employment* was four times more likely to be considered by furloughed individuals when compared to their non-furloughed counterparts (aOR 3.82 [95% CI 1.20-12.18]).

*Compressing working hours* was more likely to be considered by individuals without children in their household (8.9% compared to 3.9%, *p* = .02), and by furloughed individuals, who were nearly three times more likely to consider working compressed hours than their counterparts who had not been furloughed during the pandemic (aOR 2.91 [95% CI 1.03-8.18]).

*No employment changes* were considered by over half of those in permanent employment. In contrast, only 20.8% of those in atypical employment had not considered any employment changes at all (aOR 0.47 [95% CI 0.26-0.85]). Likewise, furloughed individuals (aOR 0.47 [95% CI 0.26-0.85]) and those with high (as opposed to low) wage precarity (aOR 0.47 [95% CI 0.25-0.88]) were significantly less likely to report not considering any change at all.

##### Employment changes considered by self-reported health

*Retraining* was more likely to be considered by those with low mental-wellbeing (33.3% compared to 21%, *p* = .02).

*Securing a permanent contract* was five times more likely to be considered by those with low mental well-being (aOR 5.49 [95% CI 1.32-22.81]).

*Becoming self-employed/freelance* was more likely to be considered by those with low mental well-(15.4% compared to 5.7%, *p* = .002). Likewise, those with limiting pre-existing conditions were four times more likely to consider self-employment than their healthier counterparts (aOR 4.00 [95% CI 1.35-11.84]).

*Retiring* was more than six times more likely to be considered by those in poorer health (aOR 6.17 [95% CI 1.29-29.52]), with 17.8% taking it into consideration (compared to 10.2% for their healthier counterparts).

## Discussion

Our study has demonstrated that when thinking about future employment, the working adult population in Wales prioritise well-paid work, within a distance close to home, that is interesting/enjoyable/rewarding, flexible, secure and with suitable working hours, and that there was little change in these key attributes during the pandemic. Although approximately half of our sample reported not considering any employment changes, more than a fifth had considered retraining since the onset of the pandemic, and many vulnerable population groups (e.g. those that were furloughed, those with atypical employment, and those with high wage precariousness) were more likely than others to consider changing their employment conditions. Comparisons across time demonstrated that having a workplace close to home became significantly more important to people as the pandemic progressed, with this being particularly true for those on permanent or fixed term contracts, and the non-furloughed. Increased time spent working from home, and the benefits it can offer for those well-equipped for home working e.g. decreased time spent commuting and increased flexibility, could account for these changes (Dyakova et al. 2021).

### The future of work and health

The extent to which health directly and indirectly relates to these employment priorities and changes is a vital consideration as employability policies. Our study showed that different population groups have different priorities and preferences for the future. Enabling equitable access to these preferred elements of work will not only make for a happier and healthier workforce, but a more productive one too (UK Government 2018). Ensuring that everyone can access work that suits their needs will help support their health.

This is particularly true for those self-reporting poorer health. Those reporting poorer general health were consistently more likely to place flexible working conditions as a priority. Furthermore, those with limiting pre-existing conditions were significantly less likely than their counterparts without such conditions to place their pay package as a priority. This suggests that other factors may take precedence for individuals living with poor health. Previous literature has highlighted how flexible working policies can help those in ill-health retain their jobs (Beatty and Joffe 2006; Holland and Collins 2018), protecting them from the negative health impacts of unemployment (van Aerden et al. 2017). Existing evidence highlights how those in ill-health and those with pre-existing conditions face barriers in obtaining and retaining work due to the challenges that their symptoms and their treatment needs present (Khan et al. 2009; Mack and Paylor 2016; van Egmond et al. 2016, 2017; Brannigan et al. 2017; Nexo et al. 2017; Booth et al. 2018; Hanson et al. 2018; Paltrinieri et al. 2018; Murray et al. 2019).

Those with limiting pre-existing conditions were 4 times more likely to consider becoming self-employed/freelance (10.1% compared to 4.9%). Those with low mental well-being also demonstrated an increased consideration of becoming self-employed/freelance (15.4% compared to 5.7% for their counterparts with better mental well-being). These findings align with prior literature, which has highlighted that turning towards self-employment is a common response for those experiencing employment difficulties arising from illness (Beatty 2012). Those with low mental well-being were also 5 times more likely to consider securing a permanent contract (15.4% compared to 5.3%), and more likely to consider retraining (33.3% compared to 21%), suggesting that these individuals want the stability that permanent contracts offer, and opportunities to develop skills that enable them to obtain alternative employment – whether this is particularly true for those whose employment conditions perpetuate their mental ill-health is a question that warrants further exploration e.g. those that experienced greater uncertainty or faced greater risks during the pandemic. With many of those with low mental well-being considering securing permanent employment, ensuring that workplaces offer mental health support that will help keep them in employment is key. This is particularly true with remote working set to be adopted more consistently beyond the COVID-19 response (e.g. Welsh Government’s aspiration to have 30% of the workforce working remotely (Welsh Government 2020a, b)). Individuals working from home during the pandemic have reported significant deteriorations to their well-being (Felstead and Reuschke 2020). Employers should provide comprehensive mental health support to their employees, whether they be home or office-based workers. The burden on mental health has been well-documented throughout the COVID-19 pandemic. The recovery period is a timely opportunity to make work-related changes that will help to ease this increased burden. Of note, our adjusted findings highlighted how those not reporting good general health were 6 times more likely than their healthier counterparts to consider entering retirement. The risk of those in ill-health exiting the labour force early due to their health-related challenges is real, and ensuring that the adaptations these individuals need are readily available will minimise the challenges they face in accessing and retaining work. Making it easier for them to access flexible working conditions, training opportunities, and alternative means of employment that offer greater autonomy (such as self-employment) and stability (as permanent contracts do) will help ensure that those in ill-health feel as able to enjoy the benefits of long working lives as their healthier counterparts.

### Employment and income related insecurity and its health burdens

Insecure work and finances can be damaging to health, with this potential being greater than ever for those placed on furlough, those with high wage precarity and those with atypical employment arrangements during the COVID-19 pandemic (Benach et al. 2014; Dyakova et al. 2021; Gray et al. 2021). These individuals are likely to require additional support during the recovery phase, with our findings suggesting that much of this will require providing additional opportunities for (and enabling access to) training, alongside improving their access to work that offers reliable hours and security within their localities. For example, those with high wage precariousness were more than twice as likely to prioritise their working hours and having a workplace close to their home. They were also twice as likely to consider retraining as their counterparts with low wage precarity. Of note, individuals with high wage precariousness (therefore experiencing financial insecurity) were more likely to fall victim to the negative economic impacts of the COVID-19 pandemic, seeing the greatest decreases in earnings, being more likely to be placed on furlough and being more likely to become unemployed (Gardiner and Slaughter 2020; Crossley et al. 2021; Gray et al. 2021). Their consideration of retraining and making employment-related changes is therefore unsurprising. In the same vein, individuals that had been placed on furlough were twice as likely to consider retraining as their counterparts who had not. Work sectors that were over-represented within the furloughed population are likely to see shifts within their labour market. Retraining creates opportunities for entering new sectors, and evidence from the US suggests that the financial strain and pandemic-induced panic y experienced by furloughed individuals within the hospitality industry during the pandemic predicted their intention to leave the hospitality industry altogether (Chen and Chen 2021). These individuals are likely to be seeking greater security and autonomy, as is reflected by the fact that furloughed individuals were four times more likely to consider securing permanent contracts, and nearly five times more likely to consider becoming self-employed/freelance. It is still too early to estimate the effects of ending the UK furlough scheme in September 2021, however concerns have been raised that furloughed individuals will be facing greater risks of unemployment following its termination (Mayhew and Anand 2020). Sectors affected by the pandemic in other ways are also seeing individuals become increasingly likely to switch sectors – healthcare workers, who worked in high-stress, high-risk environments during the pandemic, being one example (AL-Abrrow et al. 2021).

Retraining was also an attractive option for those with atypical employment contracts - half of this sub-group had considered retraining, while only a fifth of those with permanent or fixed term contracts had done the same. Atypical employment contracts are viewed to be more precarious. While they provide greater flexibility, they often offer limited stability, poorer working conditions, and often insecure hours and income (Benach et al. 2014) and with a negative impact on health (Benach et al. 2014, 2016). It is therefore of note that half of those with such contracts during the pandemic had considered accessing alternative employment through retraining. Improving access to training opportunities will support the more precariously employed to move towards work that is more conducive of their health. That being said, atypical work will remain, and efforts should also be made to ensure that the atypical work that is available is supportive of good health.

### Study implications

The COVID-19 pandemic has exacerbated societal inequalities, however the recovery phase offers the opportunity to reduce these longstanding inequalities that have become more visible during the pandemic itself. Those with low mental-wellbeing or existing mental health conditions experienced a worsening in their conditions, and increasing difficulties in accessing treatment, care and support, as did those in ill-health or with pre-existing conditions (Bu et al. 2020; Carson et al. 2020; Pierce et al. 2020; Sud et al. 2020; Zeilig et al. 2020). Precarious employment and financial insecurity are already viewed as drivers of inequalities, and the increasing uncertainty that the pandemic brought with it will have exasperated these and the associated negative impacts on both physical and mental health (Benach et al. 2014; Frattini 2017; Cheng et al. 2021). The European Parliament’s concept of “flexicurity”, introduced nearly a decade ago, remains as relevant today, with workers seeking greater flexibility and security from their work (European Parliament 2012). Those in ill-health, those experiencing financial insecurity, the furloughed and those in atypical employment considered making multiple changes to their employment conditions, and sought greater stability, more flexibility and increased autonomy. Taking these insights on board will help retain these individuals, who may already be at greater risk of leaving the labour market, in employment, particularly in light of the increased inequalities they will have faced during the pandemic. While the Welsh Government’s Employability plan and Fair Work Wales report align well with some of the key priorities for the future that are highlighted in this study (Welsh Government 2018, 2019), it is clear that more work is needed to ensure that secure, fairly rewarded work is available to all. In addition, provisions should be put in place to account for the additional training needs that might emerge as individuals consider their careers during the COVID-19 recovery and beyond. Future policies should secure targeted support that enables disproportionately affected groups and those in poorer health to pursue opportunities for retraining or entering self-employment, and ensure that employment practices give them equal access to stable, permanent employment contracts. In addition, individuals that already face health challenges should be supported in obtaining good work that is good for their health, with the provision of health support made accessible for workers, wherever they are based.

### Strengths, limitations and recommendations for future work

Our study is limited by its cross-sectional nature, whereby only associations could be calculated as opposed to causality. For example, we cannot determine whether our respondents’ were experiencing wage precariousness as a result of the pandemic, or whether it was pre-existing. However, we were able to identify changes across time within our longitudinal analyses. Second, while our study provides valuable insights about COVID-19 related changes in perspectives towards employment, they may not be reflective of individuals’ viewpoints after the removal of COVID-19 response measures (e.g. cessation of furlough, returning to the office). However, with the digital transformation of the workforce that the pandemic spurred, and the Welsh Government indicating a desire to have 30% of the Welsh workforce working remotely regularly (Welsh Government 2020a, b), many of the COVID-19 related employment impacts may remain relevant for quite some time. Thirdly, we did not account for differences across sectors in our analysis. Individuals working in certain sectors faced greater financial insecurity or increased health risks at work during the pandemic (Dyakova et al. 2021). For example, the vast majority (75%) of residential care workers reported not being able to socially distance, along with 67% of health care employees – COVID-19 related mortality was highest for social and health care workers (Office for National Statistics 2020b, 2021). Respondents working in certain sectors may have been more likely to reconsider their employment priorities or explore potential employment changes as a result of their experiences during the pandemic (as discussed for those that were furloughed within the hospitality industry and healthcare workers (Chen and Chen 2021; AL-Abrrow et al. 2021). Our findings do not capture such changes. Lastly, women and older age groups were over-represented in our sample compared to the adult workforce in Wales. The over-representation of these groups may have inflated estimates for the extent to which certain components of work were prioritised, understated the importance of others (e.g. less than 10% prioritised the availability of childcare or reliable local transport). More targeted work which allows for making longitudinal comparisons will be needed to establish whether these figures generalise to younger age groups and their intentions for their futures within the labour market in Wales, and whether any of the support recommended (e.g. mental health support, improving access to training) benefits both their employability and their health.

## Conclusion

Employment is a wider determinant of health, with the potential to generate both positive and negative effects (Burgard and Lin 2013; van Aerden et al. 2016; Julià et al. 2017). The majority of Welsh working age adults want to work close to home, with this becoming increasingly true as the pandemic progressed. Those that were furloughed, those experiencing financial insecurity, and those in ill-health all reported considering changing their employment conditions, with increasing their autonomy, flexibility and stability being a priority for these groups which may be more prone to facing insecurity within their working lives. Future policies should secure targeted support that enables these groups to pursue opportunities for retraining or entering self-employment, and ensure that employment practices give them equal access to stable, permanent employment contracts. Doing so will generate a policy environment that enables equitable access to good work that is good for health.

## Data Availability

All data produced in the present study are available upon reasonable request to the authors

## Notes

### Competing Interest Statement

The authors have declared no competing interest.

### Funding Statement

This work is supported by the National Centre for Population Health and Wellbeing Research, funded by Health and Care Research Wales.

### Author Declarations

The study was given ethical approval by the Research Ethics Committee of the National Health Service Health Research Authority, a dedicated ethics oversight body (IRAS: 282223).

