## Supplementary Material 1 for "Good work in the COVID-19 recovery: priorities and changes for the future"

### Supplementary Material 1: Questionnaire Measures

Employment priorities (asked at T1 & T2):

When looking for new or future employment opportunities, which of these are the most important to you?

Please select up to 5 options only

❑ How close the workplace is to where you live

❑ Flexible working conditions

❑ Opportunities for personal/professional development

❑ Availability of childcare

❑ Reliable local transport service

❑ Pay package (including salary, pension and benefits)

❑ Hours of work

❑ How interesting, enjoyable or rewarding the work is

❑ How well the job matches my qualifications, skills and experience

❑ Job security

❑ Other (please specify)

Employment changes (asked at T2):

Since the start of the COVID-19 pandemic (since February 2020) have you considered any of the following?

❑ Retraining to do a different job

❑ Upskilling for a promotion

❑ Securing a permanent contract

❑ Compressing your working hours

❑ Going part-time

❑ Becoming self-employed/freelance

❑ Retiring

❑ Other (please specify)

❑ None of the above

Wage precariousness computation:

Responses for three measures were used to compute wage precariousness. These included:

a. Thinking about your main job, what is your total personal income* from all sources?

❑ Less than £200 a week / less than £870 a month / less than £10,400 a year

❑ £200 to £399 a week / £870 to £1,729 a month / £10,400 to £20,799 a year

❑ £400 to £599 a week / £1,730 to £2,599 a month / £20,800 to £31,099 a year

❑ £600 to £799 a week / £2,600 to £3,459 a month / £31,100 to £41,499 a year

❑ £800 or more a week / £3,460 or more a month / £41,500 or more a year

❑ Don’t know

❑ Prefer not to say

*This is your own gross income – before any deductions like tax, national insurance, pension etc is taken off

To what extent does your income from your main job enable you to…

Always	Most of the time	Sometimes	Rarely	Never
b. cover your basic needs,

such as food, clothes,	❑	❑	❑	❑	❑

heating and housing costs?

c. cover unforeseen expenses,

e.g. urgent repair to a car,

replacement of household	❑	❑	❑	❑	❑

appliances etc?

Questions b and c were recoded onto a 0-4 scale (0 = always, 4 never). Scores for each of the three items were then divided by 12, summed, then multiplied by 4 to give a composite wage precariousness score. Scores below 1 indicate low wage precarity, scores between 1 and 1.99 indicated moderate wage precarity, and scores of 2 or above indicated high or very high wage precarity (i.e. higher financial insecurity).
